# VALIDATION OF PHLEBOTOMY PERFORMANCE METRICS DEVELOPED AS PART OF A PROFICIENCY BASED PROGRESSION INITIATIVE TO MITIGATE WRONG BLOOD IN TUBE

**DOI:** 10.1101/19010033

**Authors:** N. O’ Herlihy, S Griffin, P Henn, R. Gaffney, M.R. Cahill, A.G. Gallagher

## Abstract

**Aims:** The purpose of this study was to 1) characterise the procedure of phlebotomy, deconstruct it into its constituent parts and develop a performance metric for the purpose of training healthcare professionals in a large teaching hospital, 2) evaluate the construct validity of the phlebotomy metric and establish a proficiency benchmark.

**Method:** Using video recordings of the procedure, we defined a performance metric. This was brought to a modified Delphi meeting, where consensus was reached by an expert panel. To demonstrate construct validity, we used the metric to objectively assess the performance of novices and expert practitioners.

**Results:** A phlebotomy metric consisting of 11 phases and 77 steps was developed. The mean inter-rater reliability was 0.91 (min 0.83, max 0.95). The Expert group completed more steps of the procedure (72 Vs 69), made 69% fewer errors (19 vs 13, p = 0.014) and 300% fewer critical errors (1 Vs 4, p = 0.002) than the Novice Group.

**Conclusions:** The metrics demonstrated construct validity and the proficiency benchmark was established with a minimum observation of 69 steps, with no critical errors and no more than 13 errors in total.

## Background

Medical error is a serious patient safety issue (1) and is reported as the third leading cause of death in the US (2). Errors by doctors during the pre-analytical phase of testing i.e. before the sample is analysed in the laboratory, form a large proportion of diagnostic errors occurring in practice (3). Blood sampling is a frequently performed procedure which is prone to mistakes at several phases including identifying the patient, selecting the correct puncture site and labelling the blood specimen (4, 5). Wrong blood in tube which occurs when the blood in the bottle is not that of the patient identified on the label (6), is a critical error for patients and should be a never event. Previous research has shown that 79% of doctors report the undesirable practice of not always using wristbands for patient identification leading to a serious risk of misidentifying patients (7). Quality control in the phlebotomy process is essential. The increase in the error rate at our teaching hospital following the introduction of a new computer system and the high number of errors noted on the commencement of doctors in training in the hospital each July led to a concern regarding the safety for patients of continuing to allow doctors commence phlebotomy without adequate training. Traditionally doctors are trained according to the ‘apprenticeship’ model with a large portion of their learning taking place on sick patients. A proficiency based training programme using simulation and metric based feedback is a novel alternative. This approach has been demonstrated to be a more effective approach to training for medical procedures than traditional training models (8-10), and ensures that a large element of a doctors learning experience takes place in a simulated environment rather than in patients. The doctors must reach proficiency in the simulated environment before graduating to performing the procedure on patients.

We characterised optimal and suboptimal performance of phlebotomy to design a metric. Metrics are units used to measure performance, and break down the procedure into its constituent parts. They define how a procedure should be performed and provide a method for performance assessment (8). The metric aims to allow an individual without prior experience of the procedure to have a set of instructive steps to follow. Metric-based performance characterisation can be used to establish a proficiency benchmark which trainees must demonstrate before training progression (11).

The purpose of this study was to establish the metrics (operational definitions) required to characterise a phlebotomy procedure in doctors in training from the instruction to take blood to dispatch to the laboratory of the blood sample, to seek consensus from experts on the appropriateness of the steps and errors identified. The null hypothesis was that face and content validity for the step and error metrics derived from task deconstruction of phlebotomy procedure would not be demonstrated. Finally, we aimed to establish a proficiency benchmark to define the point at which the performance of the student has reached the mean performance of the expert group.

## Methods

### Procedure Characterisation

Procedure characterisation was performed in early 2017 over a three-month period. The metric development group consisting of a head of haematology department, a haematology consultant, medical scientists, laboratory management, laboratory information system leader, education experts, a behavioural science expert, and members of the clinical haematology team. The group met over 4 face-to face meetings to develop the metric to measure performance of the procedure of phlebotomy. Full-length videos demonstrating novice (intern) practitioners with less than 1 year experience in phlebotomy and expert practitioners with more than 5 years’ experience in performance of the phlebotomy procedure assisted in the creation and stress testing of the metric. The procedure was deconstructed and comprehensively characterised.

The phlebotomy procedure was broken down into constituent, essential, and elemental tasks necessary for the safe and effective completion of a reference approach to the performance of taking blood for laboratory testing. Particular attention was paid to the ergonomics of the procedure, preparation, patient identification and use of the computer software, which the trainees would not have practiced previously.

To stress test the metric it was subjected to an assessment of how reliably it could score a phlebotomy procedure. In a meeting with 2 Consultant Haematologists and a medical education expert the metric was used to independently score 2 videos of phlebotomy performance. Each metric was scored in a binary fashion with yes indicating the metric occurred correctly or no indicating it did not occur or occurred incorrectly. After each video differences in the scoring of each metric were reviewed and discussed. If required, operational definitions were modified, deleted or added. This process continued until the metric development group were satisfied that the metrics accurately and unambiguously characterised the specifics of the phlebotomy procedure with particular attention to blood sample labelling.

### Delphi consensus panel

The Delphi Method was chosen to test the metric using an audience of 11 experts before proceeding to validation. This method, mainly developed by Dalkey and Helmer (1963) and the Rand Corporation in the 1960’s is a widely used and accepted method for gathering data and opinion from respondents within their domain of expertise. It is designed as a group communication process, with goal-specific discussion, led by one individual or Chair (12).

At a modified Delphi meeting, an overview of the project objectives and aims was presented. An explanation of proficiency based training was given, outlining the methods and evidence behind the approach. Local and national short and long-term goals regarding improving phlebotomy proficiency to reduce incidence of Wrong Blood in Tube (WBIT) were outlined.

For the purposes of Delphi discussion, the metric was presented in ten phases. During the panel deliberations, each step was discussed and each phase was voted upon. Consensus required unanimous agreement for each step and phase. Following presentation of each phase, an opportunity was given for critical evaluation and discussion. Delphi panel members voted on whether the metric was acceptable as presented. If consensus could not be reached the metric definition was revised and a new vote conducted until a unanimous verdict reached.

### Construct Validity

To establish construct validity two groups were compared in their performance of phlebotomy. Sampling aimed to recruit at least 5 novices and 5 experts working in various specialities in the hospital including phlebotomy, nursing and medical specialities. All novices were doctors in training as the metric was primarily developed for doctors. All participants were working in Cork University Hospital at the time of the study. The expert group consisted of 2 phlebotomists, 2 haematology nurse specialists and 3 senior doctors in training. The expert healthcare professionals performed the procedure consistently in practice and were cognisant of the potential for error during blood sampling. The ‘novice’ group consisted of 5 intern doctors. Each group was instructed to perform a venepuncture on a patient. The procedure was video recorded using a Go-Pro© camera worn by the participant (first person perspective). A score of 77 would indicate that all 77 steps of the procedure had been performed correctly. If a step was missed then this was marked as an error. The videos were scored by two reviewers (director of clinical skills and a medically qualified researcher). Reviewer training consisted of a one hour meeting during which time each metric was discussed in detail. The definition of a ‘step’, ‘error’ and ‘critical error’ and how scoring should occur was clearly outlined. A full length example video was viewed and scored. Any differences in scoring methods were discussed. Finally the reviewers scored each of the videos separately. The reviewers were blinded to group status when scoring the videos. Each video was scored for ‘steps’, ‘errors’ and ‘critical errors’.

For each of the 77 steps of the procedure the numbers of ‘steps’, ‘errors’ and ‘critical errors’ were tabulated and the scores of the two reviewers were compared. The number of ‘agreements’ were tabulated (either both reviewers documented that a step was completed or both scored the step as not being completed). In addition the number of ‘disagreements’ in scoring steps was tabulated (one reviewer scored the step had been completed and the second reviewer scored that it had not been completed). The inter-rate reliability (IRR) for the steps was calculated according to the following formula: Number of agreements/ Number of agreements + Number of disagreements. An acceptable IRR is equal to or greater than 0.80 (13). Performance differences were compared for statistical significance with a one way ANOVA, using SPSS statistical package (V.24). We estimated a 26-42% difference between the experienced and novice groups based on previous studies (14).

Finally a proficiency benchmark was to be agreed to define when students could be deemed proficient to progress through training.

## Results

### Metric Development & Stress Testing the Metric

Each step in the metric was designed to be precise and unambiguous. Definitions are designed to be objective and quantitative. An effort was made to “define” rather than “describe” the performance. Steps were further characterised as “errors” or “critical errors” if they were omitted or performed incorrectly.

The 82- step metric was divided into ten phases. After 4 meetings and a final meeting to stress test the metric the procedure was characterised in its entirety. Beginning from the receipt of the order to perform phlebotomy to the delivery of the sample to the laboratory, the procedure was defined by individual steps.

### Delphi Consensus Panel

A panel of 11 experts convened in University College Cork in May 2017.

Each phase and step was discussed. The proposed metric was edited in real time and a vote was taken on the agreed consensus statement. Voting was unanimous. Consensus was reached on all phases that the metric reflected the steps necessary for safe performance of the procedure. No mandatory or essential steps had been omitted. This defined a 77 step process for safe phlebotomy, focusing on critical steps to prevent WBIT.

Modification to the metric included:

- 3 steps deleted.
- 13 steps added including the formation of a new phase “Computer”. This was noted to be vital as labels were commonly being printed prior to phlebotomy, leading to errors including mislabelling.
- 38 ording modifications for exactness or to avoid ambiguity
- 6 modifications in the order of the metric

The detail of each modification to the metric is described in the appendix. The final metric instrument consisted of 11 procedure phases and 77 procedure steps which start from the instruction to take bloods and are completed with the dispatch of the sample(s) to the laboratory (see appendix). The more serious “critical” errors were defined as those expected to either a) result in breach of patient or healthcare worker safety during the procedure itself. e.g. disposes of needle directly into the sharps bin, or b) potentially lead to a blood sampling error or wrong blood in tube event. e.g. Checks name and patient identity number on ID wrist band against the written instruction.

### Construct Validity

The mean inter-rater reliability was 0.91 (min 0.83, max 0.95). The Expert group completed more steps of the procedure (72 Vs 69), made 46% fewer errors (19 vs 13, p = 0.014) and 300% fewer critical errors (1 Vs 4, p = 0.002) than the Novice Group. This is illustrated in Figure 1. A list of the frequent errors occurring in the expert group and the novice group is displayed in Tables 1 and 2. The 11 phases of the final metric are described in Table 3.

**Table 1.** Construct validity: frequent errors which occurred in the novice group

| Number of Errors | Step Number | Description |
| --- | --- | --- |
| 6 | 22 | Checks name, MRN on ID Wrist band against the written instructions (Critical Error) |
| 6 | 38 | Performs hand hygiene before procedure |
| 6 | 47 | If using butterfly system discards the first blood tube (Critical Error) |
| 5 | 24 | Asks patient if there is any suitable vein and if one of the arms is unsuitable for venepuncture |
| 5 | 42 | If using butterfly system holds wing to insert the needle |
| 5 | 60 | Writes down patient name and DOB or MRN onto the blood tubes using a pen before leaving the patient |
| 5 | 68 | Puts closed but not locked sharps bin back where found |
| 5 | 70 | Performs hand hygiene after cleaning procedure tray |
| 5 | 54a | If using butterfly system activates the safety device during the removal process by holding down the wings and compressing sides |

**Table 2.**
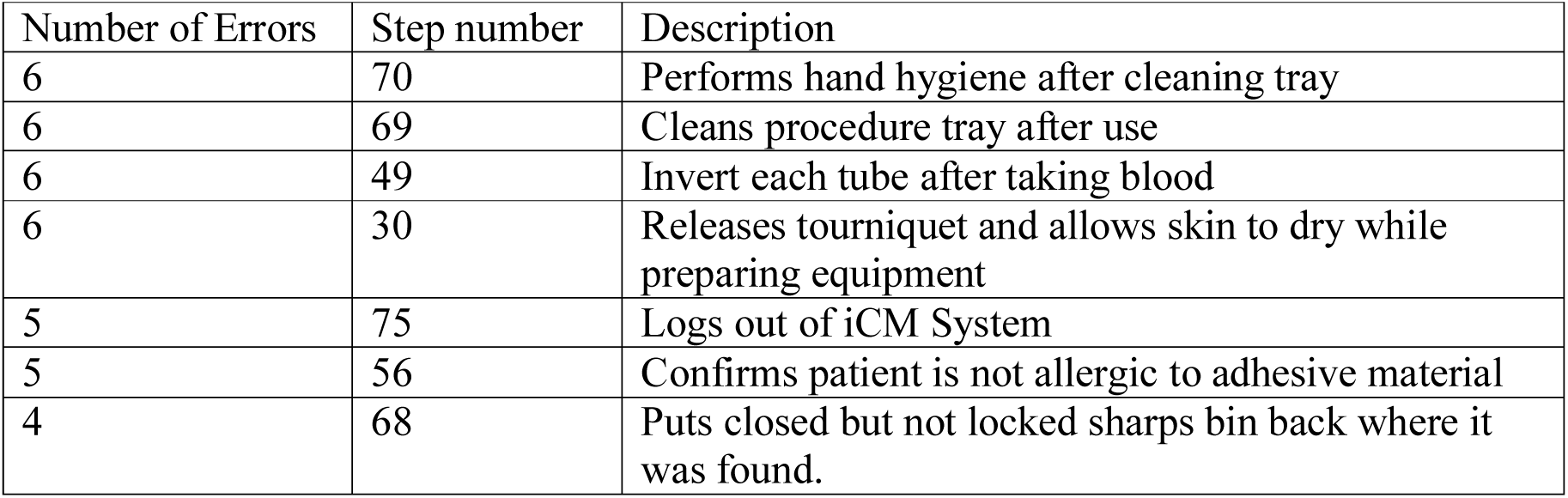
Construct validity: frequent errors which occurred in the expert group

| Number of Errors | Step number | Description |
| --- | --- | --- |
| 6 | 70 | Performs hand hygiene after cleaning tray |
| 6 | 69 | Cleans procedure tray after use |
| 6 | 49 | Invert each tube after taking blood |
| 6 | 30 | Releases tourniquet and allows skin to dry while preparing equipment |
| 5 | 75 | Logs out of iCM System |
| 5 | 56 | Confirms patient is not allergic to adhesive material |
| 4 | 68 | Puts closed but not locked sharps bin back where it was found. |

**Figure 1.**
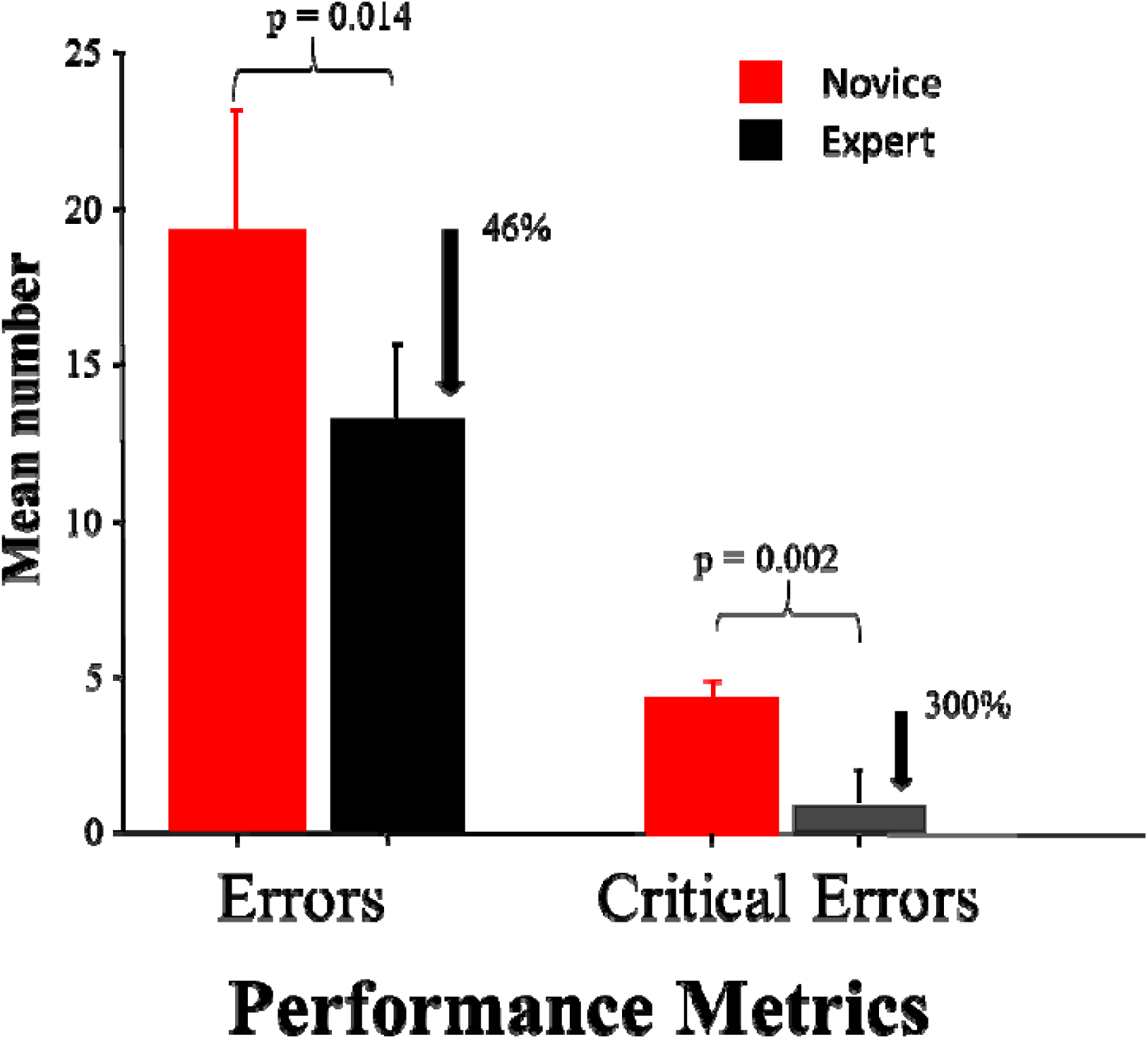
Comparison of number of error and critical errors in the Novice and Expert Groups

### Proficiency Benchmark

The proficiency benchmark was decided at a meeting of experts including, a Professor of Haematology, a Professor of Technology Enhanced Learning, a laboratory information systems leader and the medically qualified researcher. The experts had a mean of 13 errors during video recording and had completed 69 steps at least. It was decided that to reach proficiency the training programme would require the trainees to perform phlebotomy according to the metric with no more than 13 errors. Given the importance of avoiding any critical errors it was decided that no critical errors should be allowed for the trainees to reach proficiency.

## Discussion

Phlebotomy is a widely used procedure which healthcare professionals are expected to perform proficiently from their first day of work. However, due to the traditional apprenticeship model of training, many healthcare professionals do not feel adequately prepared for this skill. With the use of example videos demonstrating novice and proficient practitioners, we developed a performance metric. This was stress-tested and reviewed by a modified Delphi Meeting which accepted the metric.

The study demonstrates strong evidence of construct validity for the use of the phlebotomy metric in scoring the performance of health care practitioners while performing phlebotomy in patients. The phlebotomy metric has high inter-rater reliability indicating that it provides a precise description of the procedure with similar results when used by different reviewers. The tool can accurately distinguish between novice and expert performers of phlebotomy as the expert group had fewer errors and more importantly fewer critical errors. Previous studies have used a phlebotomy metric or checklist to evaluate VR simulation training to mannequin arm training to compare their effectiveness but evidence of construct validity for the instrument used was not clear (15-17). Guidelines exist on the correct procedure to be followed when performing phlebotomy and were developed following literature review and expert consensus alone (18-20). The construct validity of VR simulator ‘CathSim’ to distinguish between expert and novice groups has been established (21), however, the simulator does not focus on the eradication of labelling errors and there is lack evidence of concurrent validity (the closeness of the assessment to the real life environment). Our assessment tool is comprehensive outlining 11 phases and 77 steps in phlebotomy in an effort to reduce pre-analytical errors including mislabelling and wrong blood in tube errors which can be detrimental to patient safety.

### Limitations

The number of videos analysed is small with only 7 expert and 5 novice videos reviewed. The expert panel had no special qualification to determine them as experts but more years’ experience in taking blood and were aware of pitfalls in mislabelling and previous campaigns to reduce this. Only one episode of phlebotomy was observed per subject. The results may have differed if more than one procedure was observed but this was restricted by time constraints and the difficulty in recruiting subjects.

## Conclusion

The results demonstrate that the metrics can be scored reliably as indicated by the high IRR level. The metrics demonstrate construct validity and distinguished between the objectively scored performance of experienced and novice performance of phlebotomy. Based on the modified Delphi panel, the validation results and in consultation with an expert panel the proficiency benchmark was established at a minimum observation of 69 steps, with no critical errors and no more than 13 errors in total. This metric scoring instrument is available for use in training in Irish hospitals.

### Main Messages

- A validated metric for the performance of phlebotomy has been developed with a particular emphasis on avoiding mislabelling errors including wrong blood in tube.
- The metric can be used to provide a Proficiency Based Progression training programme in phlebotomy.
- A proficiency benchmark is set to ensure healthcare practitioners have reached a standard in phlebotomy equivalent to the average score of experts in the field.

### Current Research Questions

Does PBP training in phlebotomy reduce the incidence of blood sampling errors including wrong blood in tube?

Do healthcare practitioners who are provided with PBP training in phlebotomy apply the metric in clinical practice?

## Data Availability

We shall make data available to the scientific community with as few restrictions as feasible. Data will be made available by contacting the corresponding author.

---

| Phase | Procedure phase | Phase begins and ends | Definition of Phase |
| --- | --- | --- | --- |
| 1 | Introduction | Begins | Confirms patient's name and MRN OR patient's name and DOB |
|  |  | Ends | Writes down instructions including patient name, MRN, Location and types of bloods to be done and brings written instruction to bedside |
| 2 | If Crossmatch Required | Begins | Completes form using hand-writing (not patient label) |
|  |  | Ends | Completes section regarding reason for group and hold/crossmatch |
| 3 | Goes to room where equipment is kept | Begins | Decontaminate procedures tray |
|  |  | Ends | Puts last piece of equipment on tray |
| 4 | Goes to patient | Begins | Performs hand hygiene entering patient space using technique approved by WHO hand hygiene guidelines |
|  |  | Ends | Checks Name, MRN on ID Wrist Band against the written instructions (or against group and hold/crossmatch form if applicable) |
| 5 | Ergonomics of procedure | Begins | Ensure enough light to see veins |
|  |  | Ends | Releases the tourniquet and allows skin to dry |
| 6 | Prepares equipment | Begins | Positions procedure tray with sharps bin within arms' reach |
|  |  | Ends | Assembles Vacuette needle and holder with safety device off to side of black dot unless using butterfly device |
| 7 | Takes blood | Begins | Arm is positioned so that the chosen vein is pointing towards belly button of doctor (i.e. not awkward) |
|  |  | Ends | Invert each tube at least 3 times as they are taken |
| 8 | Gets ready to remove needle | Begins | Release tourniquet while last blood tube is filling before removing needle from arm |
|  |  | Ends | Disposes of needle directly into sharps bin |
| 9 | Fixes patient up after procedure and labeling of blood tube | Begins | Confirms patient is not allergic to adhesive material |
|  |  | Ends | Performs hand hygiene |
| 10 | Computer | Begins | Goes to computer, logs in and opens ICM system |
|  |  | Ends | Logs out of ICM system |
| 11 | Tidies up and sends bloods off | Begins | Puts blood tubes into sealed transport bags |
|  |  | Ends | Dispatches to laboratory in accordance with local protocol |

